# Sex differences in clinical findings among patients with coronavirus disease 2019 (COVID-19) and severe condition

**DOI:** 10.1101/2020.02.27.20027524

**Authors:** Jing Li, Yinghua Zhang, Fang Wang, Bing Liu, Hui Li, Guodong Tang, Zhigang Chang, Aihua Liu, Chunyi Fu, Jing Gao, Jing Li

## Abstract

**Objective:** To compare the sex differences in the clinical findings among patients with severe coronavirus disease 2019 (COVID-19).

**Methods:** We retrospectively collected data of 47 patients diagnosed as severe type of COVID-19 from February 8 to 22, 2020, including demographics, illness history, physical examination, laboratory test, management, and compared differences between men and women.

**Results:** Of the 47 patients, 28 (59.6%) were men. The median age was 62 years, and 30 (63.8%) had comorbidities. The initial symptoms were mainly fever (34 [72.3%]), cough (36 [76.6%]), myalgia (5 [10.6%]) and fatigue (7 [14.9%]). Procalcitonin level was higher in men than in women (0.08 vs. 0.04ng/ml, p=0.002). N-terminal-pro brain natriuretic peptide increased in 16 (57.1%) men and 5 (26.3%) women (p=0.037). Five men (17.9%) had detected positive influenza A antibody, but no women. During 2-week admission, 5 (17.9%) men and 1 (5.3%) woman were reclassified into the critical type due to deterioration. Mortality was 3.6% in men and 0 in women respectively. Four (21.1%) women and one man (3.6%) recovered and discharged from hospital.

**Conclusion:** Sex differences may exist in COVID-19 patients of severe type. Men are likely to have more complicated clinical condition and worse in-hospital outcomes as compared to women.

## Introduction

From December 2019, a pneumonia with unknown cause emerged in the Huanan seafood wholesale market, Wuhan, Hubei province. On January 3, 2020, the first complete genome of the novel β genus coronaviruses was identified in samples of bronchoalveolar lavage fluid (BALF) from a patient[1], which is different from the known 6 coronaviruses and named 2019 novel coronavirus (2019-nCoV) by WHO[2]. Soon later, the novel virus has proved to share more than 79% of same sequence with the severe acute respiratory syndrome coronavirus (SARS-CoV)[3]. Therefore, the virus is formally designated as severe acute respiratory syndrome coronavirus 2 (SARS-CoV-2) by the Coronavirus Study Group (CSG) of the International Committee on Taxonomy of Viruses[4]. The disease caused by the virus is named as coronavirus disease 2019 (COVID-19).

In the following months, SARS-CoV-2 quickly spread throughout China and even other countries[5-8]. However, the knowledge and understanding of the COVID-19 are still limited. Generally, the disease could be classified into four clinical types: mild, moderate, severe and critical pneumonia[9]. According data released by Chinese Center for Disease Control and Prevention (CDC) on February 17, 2020, a total of 44,672 cases (61.8%) has been confirmed COVID-19[10]. Among them, more than 80% are mild or moderate with relatively good short-term prognosis due to self-limiting process. However, 6,168 (13.8%) cases belong to the severe type and is more likely to develop into the critical pneumonia followed by a death rate of 49%, far beyond the average mortality of 2.3%. Although some data regarding clinical characteristics of COVID-19 has been recently revealed[5-7, 10, 11], those focusing on patients with the severe type is scarce. Moreover, since SARS-CoV-2 is highly homologous with prior SARS-CoV, it comes naturally that they may have similar clinical features, such as highly contagious infection, high lethality rate, and even sex differences[12].

In this study, we aim to compare clinical findings between men and women with severe type of COVID-19, and hope to contribute to the prevention and treatment.

## Methods

### Study population

We retrospectively collected data of 47 patients at Sino-French New Town area Tongji Hospital, Wuhan, where was supported and charged by the medical team of Beijing Hospital from February 8, 2020. Patients were all diagnosed with COVID-19 and classified into severe pneumonia according to the Diagnosis and Treatment of Pneumonia Infected by Novel Coronavirus (5th trial edition) pressed by the General Office of the National Health Commission and the General Office of the National Administration of Traditional Chinese Medicine[9]. This study was approved by the Ethics Commission of Beijing Hospital.

Laboratory confirmation of SARS-CoV-2 was done in four different institutions: the Chinese CDC, the Chinese Academy of Medical Science, Academy of Military Medical Sciences, and Wuhan Institute of Virology and Chinese Academy of Sciences. Throat-swab specimens from the upper respiratory tract that were obtained from all patients at admission were collected for extracting SARS-CoV-2 RNA. The protocol was the same as the document published recently[6]. We also examined other respiratory viruses with real-time RT-PCR, including influenza A virus (H1N1, H3N2, H7N9), influenza B virus, respiratory syncytial virus, parainfluenza virus, adenovirus, SARS coronavirus (SARS-CoV), and MERS coronavirus (MERS-CoV). Sputum or endotracheal aspirates were obtained at admission for the identification of possible causative bacteria or fungi.

Severe pneumonia met at least one of the following criteria: 1) dyspnea, respiratory frequency ≥ 30/minute, 2) blood oxygen saturation ≤93% at rest, 3) PaO2/FiO2 ratio ≤300. Critical pneumonia met at least one of the following criteria: 1) respiratory failure with mechanical ventilation, 2) septic shock, 3) transferred to intensive care unit (ICU) due to multiple organ failure.[9]

### Data collection

We obtained demographic, illness history, physical examination, laboratory test, management, and outcome data from patients’ medical records. Blood oxygen saturation was measured after oxygen therapy. Clinical outcomes were followed up by February 22, 2020.

Laboratory tests were conducted within 24 hours after admission, including a complete blood count, procalcitonin, interleukin-6, Ferritin, hypersensitive troponin I (hsTnI) and N-terminal-pro brain natriuretic peptide (NTproBNP).

### Statistical analysis

Continuous variables were expressed as median (IQR) and compared with the Mann-Whitney U test; categorical variables were expressed as number (%) and compared by χ^2^ test or Fisher’s exact test between the men and women groups. A two-sided α of less than 0.05 was considered statistically significant. Statistical analyses were done using the SPSS software (version 23) for all analyses.

## Results

From February 8 to February 11, 2020, 47 laboratory-confirmed COVID-19 patients were classified as severe pneumonia at the area, 28 (59.6%) cases were men. About 63.8% of patients had comorbidities. History of chronic obstructive pulmonary disease (COPD) was non-significant higher but hypertension and cardiovascular disease (CVD) was lower in men than in women. The initial symptoms were mainly fever, cough, myalgia and fatigue. Three men and one woman had oxygen saturation below 93% after routine nasal oxygen supply (Table 1).

**Table 1.**
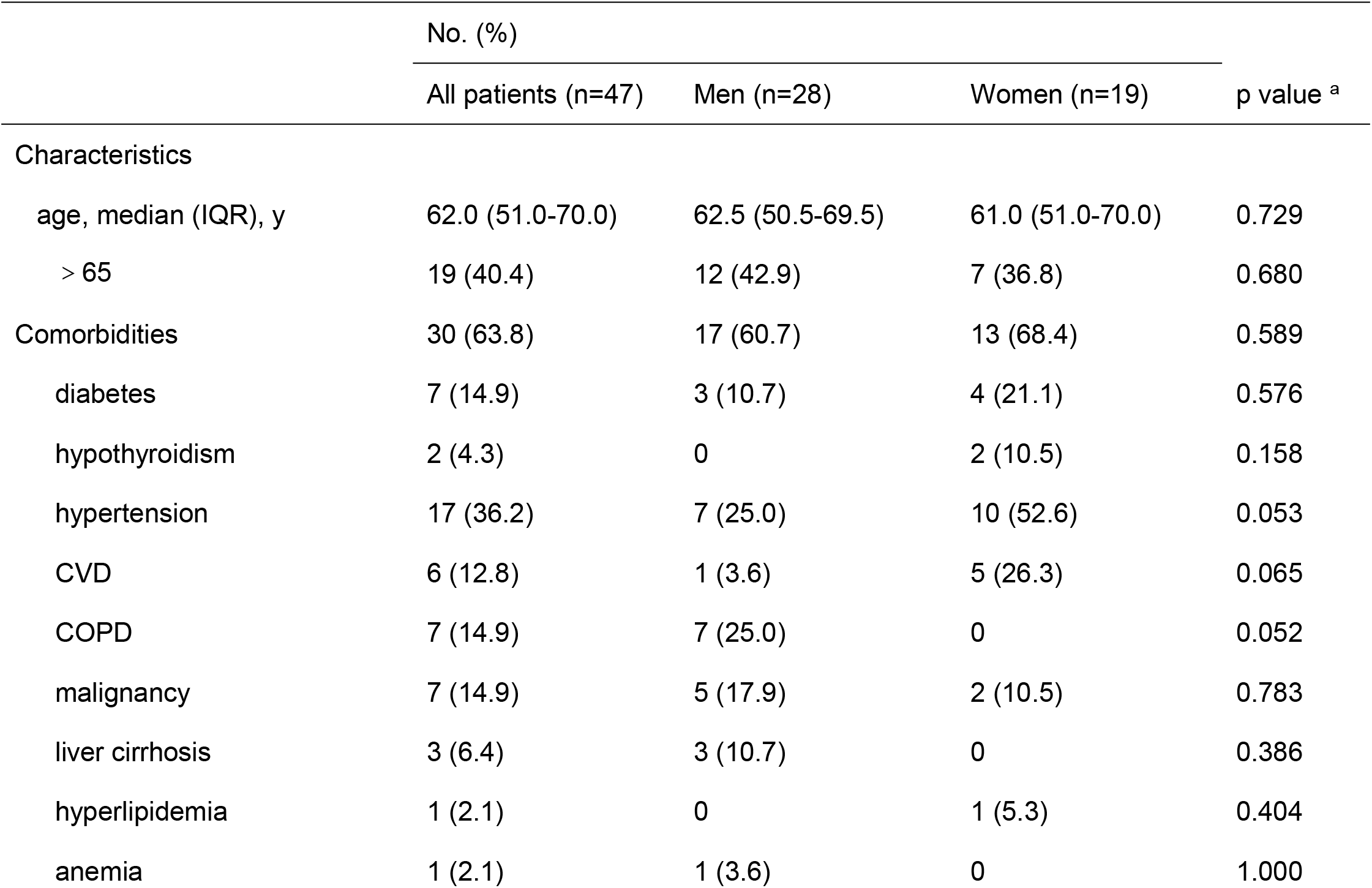
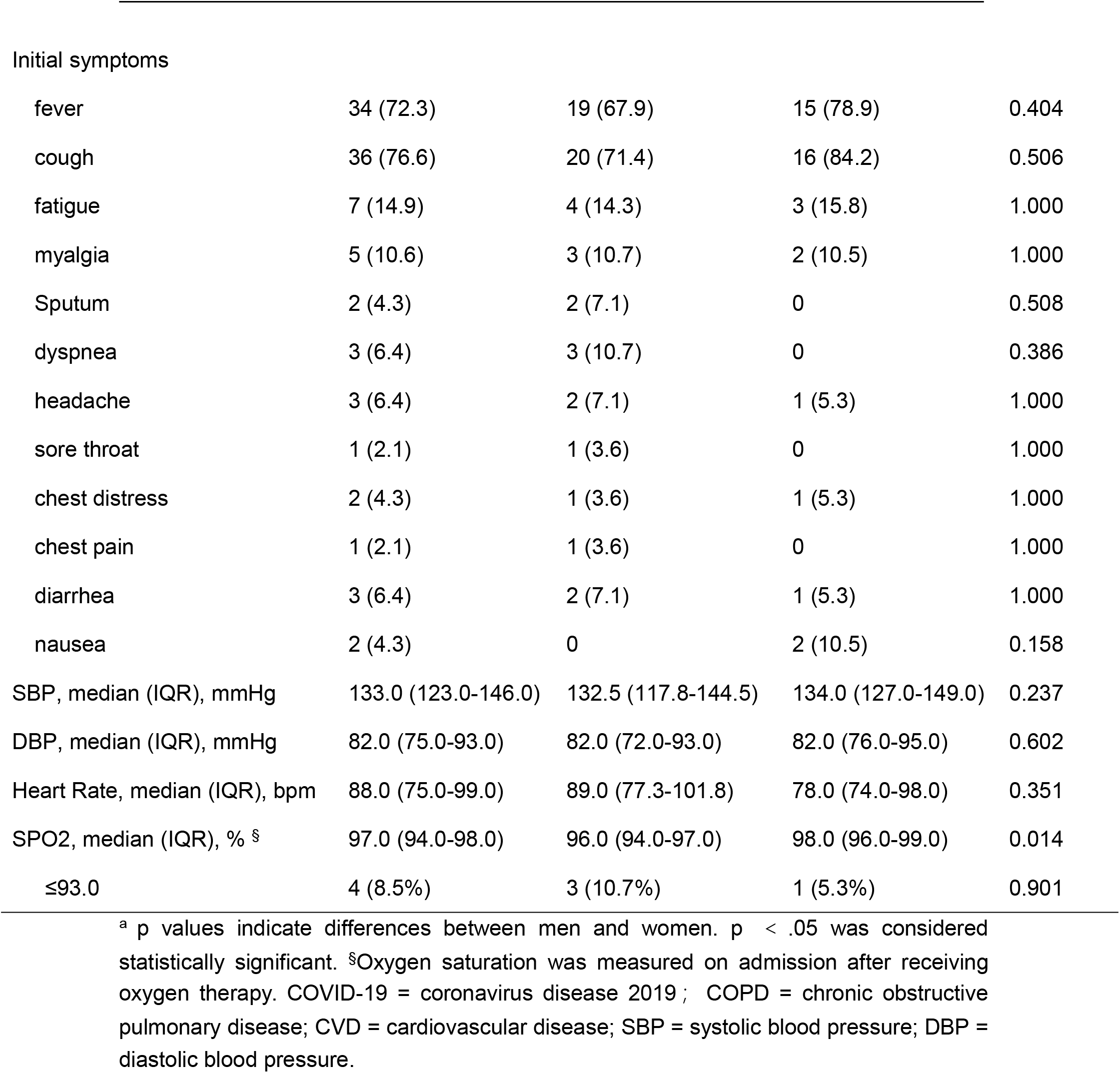
demographics and clinical characteristics of severe pneumonia patients with COVID-19.

|  | No. (%) |  |  | p value <sup>a</sup> |
| --- | --- | --- | --- | --- |
|  | All patients (n=47) | Men (n=28) | Women (n=19) |  |
| Characteristics |  |  |  |  |
| age, median (IQR), y | 62.0 (51.0-70.0) | 62.5 (50.5-69.5) | 61.0 (51.0-70.0) | 0.729 |
| > 65 | 19 (40.4) | 12 (42.9) | 7 (36.8) | 0.680 |
| Comorbidities | 30 (63.8) | 17 (60.7) | 13 (68.4) | 0.589 |
| diabetes | 7 (14.9) | 3 (10.7) | 4 (21.1) | 0.576 |
| hypothyroidism | 2 (4.3) | 0 | 2 (10.5) | 0.158 |
| hypertension | 17 (36.2) | 7 (25.0) | 10 (52.6) | 0.053 |
| CVD | 6 (12.8) | 1 (3.6) | 5 (26.3) | 0.065 |
| COPD | 7 (14.9) | 7 (25.0) | 0 | 0.052 |
| malignancy | 7 (14.9) | 5 (17.9) | 2 (10.5) | 0.783 |
| liver cirrhosis | 3 (6.4) | 3 (10.7) | 0 | 0.386 |
| hyperlipidemia | 1 (2.1) | 0 | 1 (5.3) | 0.404 |
| anemia | 1 (2.1) | 1 (3.6) | 0 | 1.000 |
| Initial symptoms |  |  |  |  |
| fever | 34 (72.3) | 19 (67.9) | 15 (78.9) | 0.404 |
| cough | 36 (76.6) | 20 (71.4) | 16 (84.2) | 0.506 |
| fatigue | 7 (14.9) | 4 (14.3) | 3 (15.8) | 1.000 |
| myalgia | 5 (10.6) | 3 (10.7) | 2 (10.5) | 1.000 |
| Sputum | 2 (4.3) | 2 (7.1) | 0 | 0.508 |
| dyspnea | 3 (6.4) | 3 (10.7) | 0 | 0.386 |
| headache | 3 (6.4) | 2 (7.1) | 1 (5.3) | 1.000 |
| sore throat | 1 (2.1) | 1 (3.6) | 0 | 1.000 |
| chest distress | 2 (4.3) | 1 (3.6) | 1 (5.3) | 1.000 |
| chest pain | 1 (2.1) | 1 (3.6) | 0 | 1.000 |
| diarrhea | 3 (6.4) | 2 (7.1) | 1 (5.3) | 1.000 |
| nausea | 2 (4.3) | 0 | 2 (10.5) | 0.158 |
| SBP, median (IQR), mmHg | 133.0 (123.0-146.0) | 132.5 (117.8-144.5) | 134.0 (127.0-149.0) | 0.237 |
| DBP, median (IQR), mmHg | 82.0 (75.0-93.0) | 82.0 (72.0-93.0) | 82.0 (76.0-95.0) | 0.602 |
| Heart Rate, median (IQR), bpm | 88.0 (75.0-99.0) | 89.0 (77.3-101.8) | 78.0 (74.0-98.0) | 0.351 |
| SPO2, median (IQR), % § | 97.0 (94.0-98.0) | 96.0 (94.0-97.0) | 98.0 (96.0-99.0) | 0.014 |
| ≤93.0 | 4 (8.5%) | 3 (10.7%) | 1 (5.3%) | 0.901 |

<sup>a</sup> p values indicate differences between men and women. $p < .05$ was considered statistically significant. <sup>§</sup>Oxygen saturation was measured on admission after receiving oxygen therapy. COVID-19 = coronavirus disease 2019 ; COPD = chronic obstructive pulmonary disease; CVD = cardiovascular disease; SBP = systolic blood pressure; DBP = diastolic blood pressure.

The serum procalcitonin and NTproBNP level were higher in men than in women. Five men had positive influenza A antibody, but no women. Antibiotic therapy was applied more in the men group than the women group (Table 2).

**Table 2.** Laboratory characteristics of severe pneumonia patients with COVID-19.

|  | Median (IQR) |  |  | p value <sup>a</sup> |
| --- | --- | --- | --- | --- |
|  | All patients (n=47) | Men (n=28) | Women (n=19) |  |
| WBC, ×10<sup>9</sup>/L | 4.7 (3.9-6.8) | 5.2 (3.6-7.6) | 4.6 (4.4-5.7) | 0.447 |
| > 10.0, No. (%) | 2 (4.3) | 2 (7.1) | 0 | 0.357 |
| < 4.0, No. (%) | 12 (25.5) | 9 (32.1) | 3 (15.8) | 0.508 |
| Neutrophil count, ×10<sup>9</sup>/L | 3.1 (2.1-5.2) | 4.1 (2.0-6.9) | 2.9 (2.5-3.4) | 0.353 |
| Lymphocyte count, ×10<sup>9</sup>/L | 1.3 (0.7-1.9) | 1.2 (0.7-2.0) | 1.3 (0.9-1.6) | 0.869 |
| ≥1.0, No. (%) | 30 (63.8) | 15 (53.6) | 15 (78.9) | 0.076 |
| Platelet count, ×10<sup>9</sup>/L | 230.0 (157.0-302.5) | 218.0 (151.3-320.8) | 230.0 (164.0-304.0) | 0.696 |
| > 300.0, No. (%) | 14 (29.8) | 8 (28.6) | 6 (31.6) | 0.825 |
| Procalcitonin, ng/ml | 0.06 (0.03-0.10) | 0.08 (0.05-0.19) | 0.04 (0.03-0.07) | 0.002 |
| increased (> 0.05), No. (%) | 25 (53.2) | 19 (67.9) | 6 (31.6) | 0.014 |
| Interleukin-6, pg/ml | 12.58 (4.05-46.66) | 17.26 (4.59-62.73) | 12.01 (2.84-33.56) | 0.402 |
| increased (≥ 7.0), No. (%) | 29 (61.7) | 18 (64.3) | 11 (57.9) | 0.658 |
| Ferritin, ng/ml | 414.0 (211.7-1131.0) | 504.0 (173.5-1256.0) | 369.0 (233.7-823.5) | 0.659 |
| hsTnI, pg/ml | 3.8 (1.9-8.5) | 4.8 (1.9-9.1) | 3.2 (1.9-6.7) | 0.230 |
| NTproBNP, pg/ml | 220.0 (63.0-338.0) | 272.5 (111.3-504.8) | 116.0 (29.0-300.0) | 0.023 |
| increased (> 241), No. (%) | 21 (44.7) | 16 (57.1) | 5 (26.3) | 0.037 |
| Influenza A antibody, No. (%) | 5 (10.6) | 5 (17.9) | 0 | 0.143 |

<sup>a</sup> p values indicate differences between men and women. p < .05 was considered statistically significant. COVID-19 = coronavirus disease 2019; WBC = white blood cell; hsTnI = hypersensitive troponin I; NTproBNP = N-terminal-pro brain natriuretic peptide;

By the end of February 22, 2020, 5 men and 1 woman deteriorated and were reclassified as critical condition. At the same period, 4 women and 1 man recovered and discharged from hospital according to the Criteria of Diagnosis and Treatment of Pneumonia Infected by Novel Coronavirus (5th trial edition). The remaining were in stable condition. (Table 3)

**Table 3.** Treatment and outcomes for severe pneumonia patients with COVID-19.

|  | No. (%) |  |  |
| --- | --- | --- | --- |
|  | All patients (n=47) | Men (n=28) | Women (n=19) |
| Treatment |  |  |  |
| antiviral therapy | 42 (89.4) | 26 (92.9) | 16 (84.2) |
| antibiotic therapy | 22 (46.8) | 15 (53.6) | 7 (36.8) |
| traditional Chinese medicine | 20 (42.6) | 11 (39.3) | 9 (47.4) |
| Clinical outcomes |  |  |  |
| discharge | 5 (10.6) | 1 (3.6) | 4 (21.1) |
| deterioration | 6 (12.8) | 5 (17.9) | 1 (5.3) |
| mechanical ventilation | 4 (8.5) | 3 (10.8) | 1 (5.3) |
| DIC | 1 (2.1) | 1 (3.6) | 0 |
| death | 1 (2.1) | 1 (3.6) | 0 |

COVID-19 = coronavirus disease 2019, DIC = disseminated intravascular coagulation.

## Discussion

In this retrospective study, we analyzed data from 47 patients with laboratory-confirmed COVID-19 of severe type. Common symptoms were similar between men and women, consistent with the recent research[6, 7, 11, 13]. Men were more likely to have prior COPD, develop secondary infections, receive complicated treatments as well as experience worse in-hospital outcomes, as compared to women.

During two-week treatment, men accounted for five of six patients (83.3%) shifting from severe to critical type. On the other side, only one man (20.0%) in five patients discharged from hospital. The similar phenomenon has also been reported in prior SARS-CoV infection in 2003. Karlberg et al.[12] analyzed 1755 SARS cases and found that men had a significantly higher mortality than women did (21.9% versus 13.2%, p < 0.0001). Leung et al.[14] showed that male sex was independently associated with a greater risk for adverse events in SARS patients. Interestingly, there is no clear evidence showing the sex difference in prognosis of influenza[15-17]. It seems that the different prognosis between men and women might be a feature for coronavirus infections.

The reason for sex difference is unknown. Some studies suggested that different outcomes between men and women could be explained by estrogen which might protect females from worse outcomes after SARS-CoV infection[12, 18]. Another possible explanation for this is that the prevalence of malignancy was higher in males. Liang et al.[19] found that in severe COVID-19 patients, those with cancer had poorer outcomes from COVID-19. In addition, more men in our study were co-infected with bacteria or influenza virus, which may somewhat result in high risk.

Rothberg et al.[20] reported that bacterial infection was a common pulmonary complication of influenza. We found that positive influenza A antibody were detected in 5 men patients but not in women. Otherwise a higher rate of increased procalcitonin was also observed in men. These findings suggested that men with severe COVID-19 is susceptible to secondary infections with virus or bacteria, resulting in higher utilization rate of advanced antiviral therapy and antibiotics. Chen et al.[11] observed that 71% of confirmed COVID-19 patients used antibiotic treatment, but no comparison between men and women. It will be clinically meaningful to further investigate why and which type of influenza virus or bacteria are more likely to co-infect with SARS-CoV-2 in men.

Another, we observed that NTproBNP but not cTnI was higher in men, suggesting more damage in heart function might exist in men after SARS-CoV-2 infection. It was reported that women were associated with less remodeling after cardiac injury, due to the protection of estrogens[21]. Our findings imply that physicians should pay more attention to cardiac function in men with severe COVID-19.

## Limitations

Our study has several limitations. First, it’s a study with small sample size, confounding factors and selection bias are inevitable. Second, the available data was limited in the early phase of SARS-CoV-2 outbreak. Third, observation period is too short to describe patients’ outcome accurately.

## Conclusion

Sex differences may exist in COVID-19 patients with severe clinical condition. Men probably have more complicated clinical status and worse in-hospital outcomes as compared to women. Further studies with large sample size are needed to verify our findings.

## Data Availability

No additional data available.

## Acknowledgments

The study was funded by the Beijing Municipal Natural Science Foundation (No.7192078). We thank all patients involved in this study.

## Conflict of interest

All authors have nothing to declare.

## Notes

### Competing Interest Statement

The authors have declared no competing interest.

### Funding Statement

The study was funded by the Beijing Municipal Natural Science Foundation (No.7192078). The funder was not involved in the collection, analysis, or interpretation of data, or in the writing or submitting of this report.

